# Can AI-generated clinical vignettes in Japanese be used medically and linguistically?

**DOI:** 10.1101/2024.02.28.24303173

**Authors:** Yasutaka Yanagita, Daiki Yokokawa, Shun Uchida, Yu Li, Takanori Uehara, Masatomi Ikusaka

## Abstract

**Background:** Creating clinical vignettes requires considerable effort. Recent developments in generative artificial intelligence (AI) for natural language processing have been remarkable and may allow for the easy and immediate creation of diverse clinical vignettes.

**Objective:** In this study, we evaluated the medical accuracy and grammatical correctness of AI-generated clinical vignettes in Japanese and verified their usefulness.

**Methods:** Clinical vignettes in Japanese were created using the generative AI model GPT-4-0613. The input prompts for the clinical vignettes specified the following seven elements: 1) age, 2) sex, 3) chief complaint and time course since onset, 4) physical findings, 5) examination results, 6) diagnosis, and 7) treatment course. The list of diseases integrated into the vignettes was based on 202 cases considered in the management of diseases and symptoms in Japan’s Primary Care Physicians Training Program. The clinical vignettes were evaluated for medical and Japanese-language accuracy by three physicians using a five-point scale. A total score of 13 points or above was defined as “sufficiently beneficial and immediately usable with minor revisions,” a score between 10 and 12 points was defined as “partly insufficient and in need of modifications,” and a score of 9 points or below was defined as “insufficient.”

**Results:** Regarding medical accuracy, of the 202 clinical vignettes, 118 scored 13 points or above, 78 scored between 10 and 12 points, and 6 scored 9 points or below. Regarding Japanese-language accuracy, 142 vignettes scored 13 points or above, 56 scored between 10 and 12 points, and 4 scored 9 points or below. Overall, 97% (196/202) of vignettes available with some modifications.

**Conclusions:** Overall, 97% of the clinical vignettes proved practically useful, based on confirmation and revision by Japanese medical physicians. Given the significant effort required by physicians to create vignettes without AI assistance, the use of GPT is expected to greatly optimize this process.

## Introduction

Clinical vignettes are clinical education cases used to measure learners’ knowledge and clinical reasoning abilities. They are designed to assess the skills needed by learners to perform tasks required for diagnosis and treatment, such as medical history taking, physical examination, test ordering, and management.^1^ They are used by educators, demographers, and health service researchers; further, they have been widely adopted to measure processes in various clinical settings.^2^ Each simulated case in clinical vignettes contains realistic clinical details, allowing for the presentation of the same clinical scenarios to multiple physicians.^3^ They also enable a concrete understanding of patient cases, even for less experienced learners, and serve as content for doctors’ continuing professional development. Therefore, the use of clinical vignettes is expected to help mitigate the disparities in physicians’ experience levels and clinical experiences across different regions.^4^

However, creating clinical vignettes requires considerable effort. These models are typically constructed using detailed clinical information based on actual clinical practice, assuming a single patient. Even for clinical vignettes of the same disease, the frequency of symptoms, course, test results, and treatment effects are not uniform; therefore, the number of vignettes that reflect real clinical practice can become immense. Therefore, it is necessary to create vignettes that reflect the intentions of educators and users, tailored to the level of learners and the abilities being measured.

OpenAI (San Francisco, California, USA) has released a generative artificial intelligence (AI) model, the Generative Pretrained Transformer (GPT), which can utilize natural language to answer any question,^5^ and its usefulness is being increasingly recognized. GPT can understand the meaning of questions posed by users and provide answers to questions on a wide range of topics across various domains, including general knowledge, programming, mathematics, science, philosophy, etc. Depending on the prompts entered, it can also instantly generate novels or essays.^6^ This generative AI is available to anyone and its technological development is advancing rapidly. Consequently, more powerful versions of GPT have been developed, significantly improving the accuracy of its outputs.^7^ In the field of clinical medicine and medical research, GPT has been reported to be capable of answering questions related to the medical licensing exam and specialist-level knowledge.^8–10^ It is also capable of clinical simulations,^11^ while its application in medical education is currently being discussed.

Generative AI also holds potential as a virtual educational assistant.^12^ Using generative AI, there is the possibility of creating high-quality clinical vignettes can be created with minimal effort and in a short amount of time. However, caution is necessary because there is generally no guarantee that the content output of generative AI is based on evidence.^13^ Moreover, the accuracy AI-generated clinical vignettes can only be assessed through a peer review process conducted by physicians. Furthermore, while the GPT primarily provides services in English, it has become clear that the accuracy of outputs in Japanese is not always satisfactory,^14^ necessitating evaluation by Japanese speakers for both medical and Japanese-language accuracy. Considering our previous research, in which differential diagnosis lists and illness scripts were created based on medical information,^15^ it is expected that the creation of clinical vignettes is similarly feasible. However, no studies have aimed to create and evaluate AI-generated clinical vignettes in Japanese. Therefore, this study aimed to evaluate the medical and Japanese-language accuracy of AI-generated clinical vignettes.

## Methods

A cross-sectional analysis was conducted in which clinical vignettes were created using the generative AI model known as GPT. The vignettes were evaluated by three Japanese physicians using a five-point scale.

### Production environment

We performed the analysis on a terminal using the operating system Ubuntu 20.04.2 LTS; its CPU was an AMD Epyc 7402p 24-core processor and its primary memory was 256 GB. An A100 graphics accelerator (NVIDIA) with 40 GB of RAM was used for the computation. We used Python (3.6.9) and Openai (0.27.8), which are OpenAI Python libraries. The API utilized GPT-4-0613 to create a clinical vignette on June 13, 2023. GPT is an AI language model based on the generative pretrained transformer architecture, a type of neural network designed for natural language processing tasks, developed by OpenAI. It generates on-the-spot responses based on the likelihood of the next word using the relationships between words within the neural network learned during training^16^.

### Selection of target diseases for creating clinical vignettes

From the viewpoint of the diseases that beginner physicians should learn about, disease groups in the primary care domain were selected as the targets for creating clinical vignettes in this study. Based on the diseases and signs targeted in family physician training (which aims to teach medical professionals how to manage general diseases and conditions in coordination with other specialists and healthcare professionals as needed^17^), 202 cases were selected following discussions among three physicians—one board certified member of the Japanese Japanese Society of Internal Medicine (YY) and two Japan Primary Care association certified family physicians (SU and DY) (Supplement). In addition, three diseases (Parkinson’s disease, dermatomyositis, and infectious endocarditis) that were entered as model clinical vignettes during the prompt phase were excluded.

### Prompts entered into GPT

The content input into the GPT was made comprehensible to the AI by referencing prompt engineering techniques. In order to provide the user with the desired output, the input was kept brief. The Act As Pattern prompt was modified to assign the AI the role of an author writing case summaries for physicians.^18^ Furthermore, by referencing abstracts typically presented at general medical conferences, instructions were given to output each clinical vignette in Japanese within 700 characters. The items required to fulfill the vignette format were entered in the following order: 1) age, 2) sex, 3) chief complaint and time course since onset, 4) physical findings, 5) test results, 6) diagnosis, and 7) treatment course. Additional regulations were noted, such as not outputting reference values for blood tests or discussions and limiting the use of pharmaceuticals in Japan. Finally, three model clinical vignettes created by AI and revised by physicians were entered (infective endocarditis, dermatomyositis, and Parkinson’s disease).

### Evaluation of the clinical vignettes

Regarding the verification process for the output cases, an internal medicine physicians (YY) initially confirmed whether the seven items specified in the prompt for the clinical vignette format—(1) age, 2) sex, 3) chief complaint, course since onset, 4) physical findings, 5) test results, 6) diagnosis, and 7) treatment course—were present in the output. Subsequently, two family physicians (DY and SU) participated in the evaluation process and created a three-member team assessment. There were two evaluation items: medical accuracy and Japanese-language accuracy. Each evaluator assigned a score on a five-point scale ranging from 5 (“very useful, no additional modifications needed”) to 1 (“not at all useful, general modifications needed”). The five-point scores for each of the two criteria from the three evaluators were integrated to form a composite score of 15 points. A composite score of 13 points or above was defined as “sufficiently beneficial and immediately usable with minor revisions,” a score of 10–12 points was considered was defined as “partly insufficient and in need of modifications,” and a scores of 9 points or below was defined as “insufficient.”

### Ethical considerations

This study did not involve human or animal participants and ethical approval was not required.

## Results

Using GPT-4-0613, clinical vignettes for 202 diseases were created. In all the output clinical vignettes, the seven items specified in the input prompts were output without any omissions. Regarding medical accuracy, 118 vignettes (58.4%) scored 13 points or above, 78 (38.6%) scored between 10 and 12 points, and 6 (3.0%) scored 9 points or below (Figure 1). Overall, 97% (196/202) of vignettes available with some modifications. Regarding Japanese-language accuracy, 142 vignettes (70%) scored 13 points or above, 56 (27.7%) scored between 10 and 12 points, and 4 (2.0%) scored 9 points or below. Furthermore, a moderate positive correlation was observed between medical accuracy and Japanese-language accuracy (correlation coefficient = 0.43) (Figure 2). The clinical vignettes with high and low medical accuracy evaluations are shown in Figures 3 and 4, respectively.

**Figure 1.**
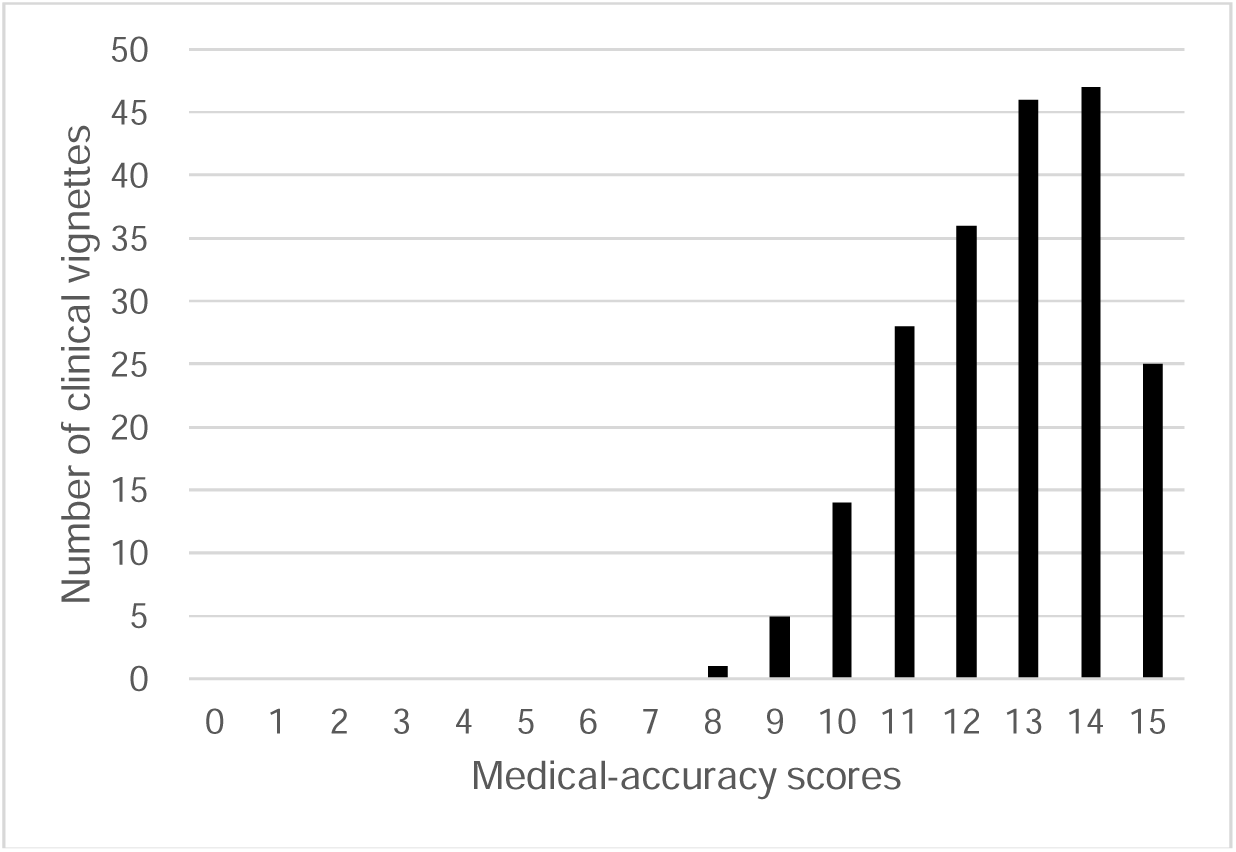
Evaluation of medical accuracy

**Figure 2.**
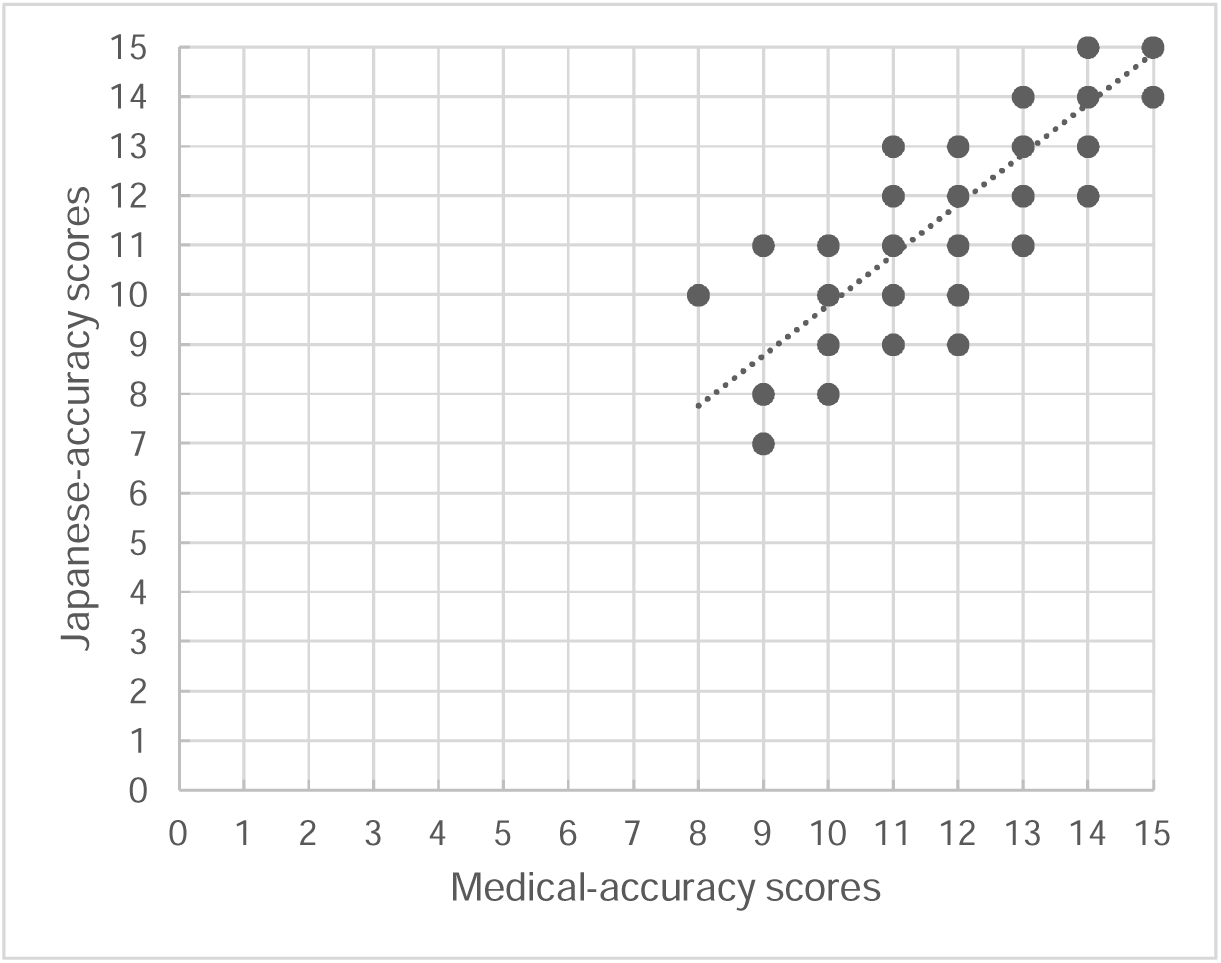
Correlation between the evaluation of medical and Japanese-language accuracy

**Figure 3.** Clinical vignette with high medical accuracy Case) Depression A 45-year-old woman began experiencing a depressive mood, loss of interest, and poor appetite three months ago, with sleep disturbances starting two months prior. Her chief complaint was a pervasive deep feeling of melancholy throughout the day, making it difficult to go to work. She also had no energy for anything, experienced low self-esteem, and could not find hope for the future. Physical examination showed a gloomy facial expression and slow responses. She had lost weight and was eating very little. Blood tests did not reveal any abnormalities related to diseases with a high prevalence of concomitant symptoms, such as hyperthyroidism or anemia. No significant lesions were found on a head CT scan. From the medical history and findings, she was diagnosed with Major Depressive Disorder. For treatment, she was started on 20mg daily of the selective serotonin reuptake inhibitor (SSRI) fluoxetine, along with cognitive-behavioral therapy. After one month of treatment, sleep disturbances and poor appetite improved, and subsequently, her mood disorder also lessened. However, as the low self-esteem persisted after three months, increasing the fluoxetine dose to 40mg is being considered. Cognitive-behavioral therapy is also being continued.

**Figure 4.** Clinical vignette with low medical accuracy Case) Primary amenorrhea Twenty-five years old, female. She had been without menstruation for 6 months prior to the initial visit and had subjective symptoms of breast tenderness and depression. She had a history of sexual intercourse and used a condom as a contraceptive method. There were no major lifestyle changes, such as weight gain, appetite changes, or changes in exercise habits. Physical examination revealed no abnormalities in body size, skin, or thyroid gland after detailed observation. The breasts were checked for the presence of non-papillary secretions, which were confirmed as normal. Blood tests showed that blood hormone levels, including luteinizing hormone (LH), follicle stimulating hormone (FSH), prolactin, thyroid stimulating hormone (TSH), and free T4 were within normal ranges, and human chorionic gonadotropin (hCG) was negative. Gynecological examination revealed no abnormal findings in the uterus or ovaries. Based on these findings, a diagnosis of primary amenorrhea was made. Estrogen and progesterone replacement therapy was initiated. Two cycles of Esclvere (Esmya) at 5 mg once daily for 28 days were prescribed. Vitamin D and calcium were additionally supplemented once daily. Menstruation resumed approximately 3 months after the start of treatment, and breast tension and depressed mood also improved. She was followed up regularly thereafter and instructed to continue using estrogen and progesterone according to her menstrual cycle. Her blood hormone levels remained within the normal range, and her findings remained stable. At present, no signs of recurrence have been observed.

Focusing primarily on clinical vignettes that were subject to deductions for each item, three physicians (YY, US, and DY) engaged in comprehensive discussions to identify the areas for improvement. The specific inadequacies identified concerning the medical and Japanese-language accuracies of the clinical vignettes are presented below.

1) Examples of vignettes with lower medical accuracy:

1. Insufficient information: No history of smoking in the case of lower limb arteriosclerosis. / In the case of aortic valve stenosis, the finding was of mitral valve murmur, not aortic valve murmur.
2. Inappropriate tests: For deep vein thrombosis, an MRI was performed without an echo or contrast-enhanced CT.
3. Inappropriate diagnostic basis: The diagnosis of impetigo based on the culture of pus.
4. Inappropriate evaluation: Blood test values were evaluated as normal despite being abnormal / A temperature of 36.8°C evaluated as a low-grade fever.
5. Inappropriate treatment: Use of anticoagulants for the treatment of cerebral hemorrhage / Prescription of antibiotics for a cold / Treatment of RSV infection with ceftriaxone.
6. Inappropriate medication dosing: Oral bisphosphonate alendronate 70mg/week for osteoporosis treatment is not an approved dosage under Japanese insurance.
7. Inappropriate management: Regular follow-up checks at 1 month, 3 months, and 6 months post-mumps for complications like meningitis / Recommending urology follow-up for the treatment of compression fracture.

2) Examples of vignettes with lower Japanese-language accuracy:

1. Grammatical errors in Japanese.
2. Unnatural medical expressions.
3. Output of non-existent disease names or drug names.
4. Instances of word repetition.
5. Use of katakana for English terms.
6. Output of Chinese or Korean language in phrases.

## Discussion

By specifying seven input prompts, 202 clinical vignettes were created for common diseases and conditions in primary care. Of these, 118 (58.4%) were judged by all three physicians to have high medical accuracy. When including those deemed “partly insufficient and in need of modifications,” the total usable clinical vignettes amounted to 196 (97%). However, critical issues were also extracted, such as nonexistent disease names and inappropriate treatment. Furthermore, 142 (70%) were judged to have high Japanese-language accuracy. It is clear that GPT can create highly accurate Japanese-language clinical vignettes of up to 700 characters if revised by Japanese physicians. Creating clinical vignettes conventionally is an effort-intensive process because it requires physicians to consult numerous reference books to compile all the necessary clinical information. Furthermore, despite the use of various references, vignettes, being a variant of the case study method, tend to reflect the personal experiences of their creators. On the other hand, vignettes generated by AI could be standard and representative, potentially making them suitable for novice learners. Additionally, GPT is capable of generating vignettes in a short amount of time. Clinical vignettes have been proven to be practical in the past,^19^ and medical education using clinical vignettes created by the GPT could open up new possibilities for lifelong education, not just for students.

However, the greatest challenge in developing AI-generated clinical vignettes is ensuring high medical accuracy. Regarding the vignettes’ medical accuracy, critical issues such as insufficient information, missing physical examination data, inappropriate tests, inadequate assessments, prescription of unnecessary medication, dosages exceeding the recommended amounts, and non-recommended management were observed. In particular, caution is necessary with outputs that include dosages adopted abroad that differ from those used in Japan, prescriptions of contraindicated drugs, and those inappropriate from the standpoint of antimicrobial resistance. These outputs could lead to medical errors if used unmodified by healthcare workers with insufficient knowledge in the relevant field because they may not recognize these mistakes. Another inappropriate output was that some clinical vignettes suggested regular follow-up observations for conditions that did not require them, even recommending checkups every few months. While the reasoning process of the generative AI’s output is a black box and therefore speculative, the influence of the three sample-case inputs (Parkinson’s disease, dermatomyositis, and infectious endocarditis), which require long-term observation, cannot be ruled out. Additionally, there was one case that presented an unknown disease name. Non-existent drug names were occasionally observed.

This study evaluated 202 AI-generated clinical vignettes created using a single input. Although only one input-output cycle was performed in this study, it is possible that better vignettes could be produced through repeated similar inputs.^20^ At present, prompt modification is considered the most accurate method to improve the output within a correctable range. By modifying the prompts, it is possible to suppress unpredictable outputs, align the output with the instructions, and refine the text.^21^ Moreover, there is a significant possibility of creating higher accuracy and more desirable cases for users by instructing the generative AI on specific points to be revised for each individual case. When using this generative AI and the output clinical vignettes for medical education or other purposes, it is possible to create clinical vignettes with less effort by carefully evaluating and modifying the appropriateness of the test selection and dosage of therapeutic drugs while confirming the patient’s medical history.

Regarding the vignettes’ Japanese-language accuracy, there were instances of grammatical mistakes and unnatural expressions. One reason for this is that the GPT is trained on an English database; hence, translations into other languages, including Japanese, have limitations.^22^ Moreover, Japanese belongs to a different language family tree than languages derived from Latin, such as English, which might pose an affinity issue for English-based databases. Moreover, natural language processing technologies are imperfect. However, if a native Japanese speaker reads the output, unnatural points can be easily identified within a range that is straightforward to correct. Among the clinical vignettes output in this study, three were produced in Chinese or Korean. Although it is difficult to control the output of other languages using prompts, it is easy to correct this issue if the vignettes are reviewed by a Japanese speaker. These outputs are believed to occur because the GPT relies on publicly available information that it has learned. As GPT’s learning of medical information in Japanese progresses, its accuracy is likely to improve.^23^

As evident from the clinical vignettes created in this study, considering the utility of medical information being instantly available through generative AI, it is expected that the technology of this generative AI will be increasingly applied to medical education and healthcare in the future.^24^ Although there remain valid concerns about copyright issues and the medical accuracy of the content produced by generative AI,^25^ if these technologies are used with due consideration, their application has nearly boundless potential. In particular, within the scope of these technologies’ use for medical education, it is possible for instructors to censor the output, which allows its use in providing to students to be sufficiently permissible.^26^

In sum, this study confirms it is possible to create and validate clinical vignettes of rare diseases and atypical cases of high-frequency diseases using generative AI. Moreover, the generation of images and videos using AI is now also feasible^27;^ this opens up the possibility for creating clinical vignettes containing images of X-rays, computed tomography, magnetic resonance imaging, and electrocardiography, which could further enrich the content of AI-generated vignettes and make them more educational.

## Limitations

This study had three limitations. First, the version of GPT used in this study was GPT-4-0613, and the evaluation was based on the outputs generated as of June 13, 2023. Because these technologies’ accuracy may change with future updates, regular assessments are necessary.

Second, there were no clear criteria for evaluating the clinical vignettes in this study, which were assessed based on the subjective viewpoints of three general physicians; thus, the results may differ if evaluated by physicians from other specialties. The third limitation is that, by design, the generative AI cannot create reproducible outputs, which is considered a drawback. In addition, there is no standardized method for inputting prompts, and accuracy may be affected by the input content. The extent to which input content affects the results has not been verified and remains a challenge for future research.

## Conclusion

The use of a generative AI, GPT, in conjunction with corrections by Japanese physicians enabled the creation of Japanese-language clinical vignettes with 97% medical and linguistic accuracy. Creating clinical vignettes conventionally is an effort-intensive process; therefore, GPT usage can be expected to significantly reduce the required time investment. Additionally, the task of making corrections to the output can be used in various settings, including continuing medical education for physicians and AI-assisted medical education for medical students. With further advances in AI technology, improvements in the accuracy of AI-generated clinical vignettes are expected.

## Author contributions

YY, DY, and MI designed and coordinated this study. YY, DY, SU, and UT performed data analysis and interpretation. YY, DY and MI drafted the manuscript. SU, YL, TU, and MI revised the manuscript for intellectual content. All authors read and approved the final manuscript and agreed to be accountable for all aspects of the work, ensuring that questions related to the accuracy or integrity of any part of the work were appropriately investigated and resolved.

## Data Availability

The data regarding the results of this study are available from the corresponding author (YY) upon reasonable request.

## Supplement

List of 202 targeted diseases.

